# Improving risk communication: a pilot randomised-control trial assessing the impact of visual aids for interventional consent

**DOI:** 10.1101/2023.06.04.23290923

**Authors:** Despoina Chatzopoulou, Arif Hanafi Bin Jalal, Danail Stoyanov, Hani J Marcus, Anand S Pandit

## Abstract

**Introduction:** Informed consent is a fundamental component in the work-up for surgical procedures. Our study aims to assess the impact of visual tools in the informed consent process of a common medical and neurosurgical procedure: a lumbar puncture.

**Methods:** Participants were healthy adults without any underlying cognitive impairment, and they were randomized to complete a questionnaire containing either the control video or intervention video. Both videos contained identical audio narration, however the intervention video included additional visual aids such as anatomical diagrams, icon arrays and Paling scales. Outcome measure included knowledge of the procedure (including recall of numerical risks), 5-point Likert scale questions, usability as measured by the System Usability Scale, acceptability as measured by the Acceptability of Intervention Measure and appropriateness as measured by the Intervention Appropriateness Measure.

**Results:** In total, 52 participants were randomized to the control (n = 25) or intervention (n=27) group. There was no statistical difference in numerical risk recall, understanding of the procedure or its benefits. The intervention group reported better understanding of the risks (p = 0.05), felt they could more easily better explain the risks to others (p = 0.01), and felt less overwhelmed with information (p = 0.03). The intervention group was also rated as more acceptable (p = 0.02).

**Conclusion:** Our pilot study tentatively demonstrates that visual risk communication adjuncts offer several advantages over traditionally obtained surgical consent without being inferior in understanding of the procedure or recall of numerical risks.

## Introduction

Informed consent is a fundamental component in the work-up for surgical procedures. Statistical risk information pertaining to the procedure is by nature probabilistic and challenging to communicate, especially to those with poor numerical literacy (1). Our pilot study demonstrates the use of visual statistical risk communication adjuncts in a simulated procedural consent of a lumbar puncture. We analyse participants’ understanding of the procedure, but also more specifically, their understanding of procedural complications, associated numerical probability and attitudes related to surgical consent.

## Methods

Adults above 18 years old were recruited among staff and students at our institution following study approval by the local institutional ethics committee and retrospective registration on ClinicalTrials.gov (NCT05717465). The educational background of the participants varied from secondary level of education to postdoctoral level of studies within and outside the medical field (Table 1). Exclusion criteria included individuals with prior experience of the procedure, lacking capacity to consent or having an underlying cognitive impairment.

**Table 1.** Table of Demographics between Control Group and Visual Aids Group. MWU = Mann-Whitney U test, FE=Fischer’s exact test, χ2=Chi-squared test

|  | **Visual Aid**<br>**Group (n=25)** | **Control**<br>**Group (n=27)** |  |
| --- | --- | --- | --- |
| **Sex** |  |  | p-value = 0.48 (FE) |
| Male | 12 | 11 |  |
| Female | 12 | 16 |  |
| Prefer not to say | 1 | 0 |  |
| **Age** |  |  | p-value = 0.32 (MWU) |
| Age (median and age range) | 23 (19-56) | 22 (19-28) |  |
| **Professional status** |  |  | p-value = 0.34 (FE) |
| Student on another course | 10 | 12 |  |
| Medical student | 6 | 10 |  |
| Professional in non-healthcare related field | 9 | 5 |  |
| **Completed education** |  |  | p-value = 0.07 (FE) |
| Secondary school | 5 | 4 |  |
| Diploma / College certificate | 3 | 3 |  |
| Bachelor's / Master's degree | 12 | 20 |  |
| Doctoral / PhD degree or higher | 5 | 0 |  |
| **Median familiarity with procedure** | 1.96 | 1.96 | p-value = 0.96 (MWU) |
| **Method of completion** | | | p-value = 0.50 ($\chi^2$) |
| Laptop | 18 | 16 |  |
| Mobile device (phone, tablet) | 7 | 11 |  |

Participants were randomly allocated to one of two groups and sample size calculations were performed, with a minimum of 25 participants in each group required for a significant difference of 0.4 points between groups based on a 5-point Likert scale (Cohen’s *d* = 0.8, alpha = 0.05, power = 0.80, GPower v=3.1). While both groups received identical medical information content regarding hypothetical clinical scenarios in which an intervention was consented for, the means of consent differed (Figure 1). The control group (Group A) had the same risks verbally explained without any aids as is typical for a pre-operative surgical consent. In both groups, the consent and statistical information was pre-recorded to ensure homogeneity and a written consent form was presented at the end of the video (Supplementary Methods). The trial group (Group B) received the statistical information in the form of visual aids - anatomy and Paling scales (Supplementary Figure 1,2) (2). The video recording was made with the use of Vyond (Vyond, San Mateo, Ca), a cloud-based video animation tool. The same voice recording was used in both groups, ensuring the content of the medical information is identical in both groups, with the addition of audio-visual tools only in the trial group.

Participants completed anonymised questionnaires relating to how they recalled and perceived statistical risks, on their understanding of the procedure and how usable, acceptable, and appropriate their consent method was, using validated scales (Supplementary Methods) (3). All statistical analysis was performed in RStudio (RStudio Team, Boston, MA). Normality of the data was assessed using the Kolmogorov-Smirnov test, and by visually inspecting the distribution. If data was non-parametric, the Mann-Whitney U test was used to assess the differences between the control and intervention groups, with a p-value < 0.05 being considered as significant.

## Results

52 participants completed the questionnaire, and no statistically significant differences were found in the age, sex, professional status, understanding of, and prior familiarity with the procedure between groups, although the trial group had a trending higher level of overall literacy (Table 1). There was no statistical difference in numerical risk recall, and those in the intervention (visually-enhanced consent) group were not inferior in their subjective understanding of the procedural benefits (p = 0.29) [Table 2]. However, the intervention group seemed to have a better understanding of the risks (p = 0.05), they thought they could better explain easier the risks to others (p<0.01), and they seemed to feel less overwhelmed with information (p = 0.03). Furthermore, the enhanced consent process was found by participants to be significantly more acceptable (p = 0.03). It showed a trend towards greater appropriateness (p = 0.06) and it appeared to have ‘good’ usability (median SUS = 76.4), although this also did not reach statistical significance (p = 0.06).

**Table 2.**
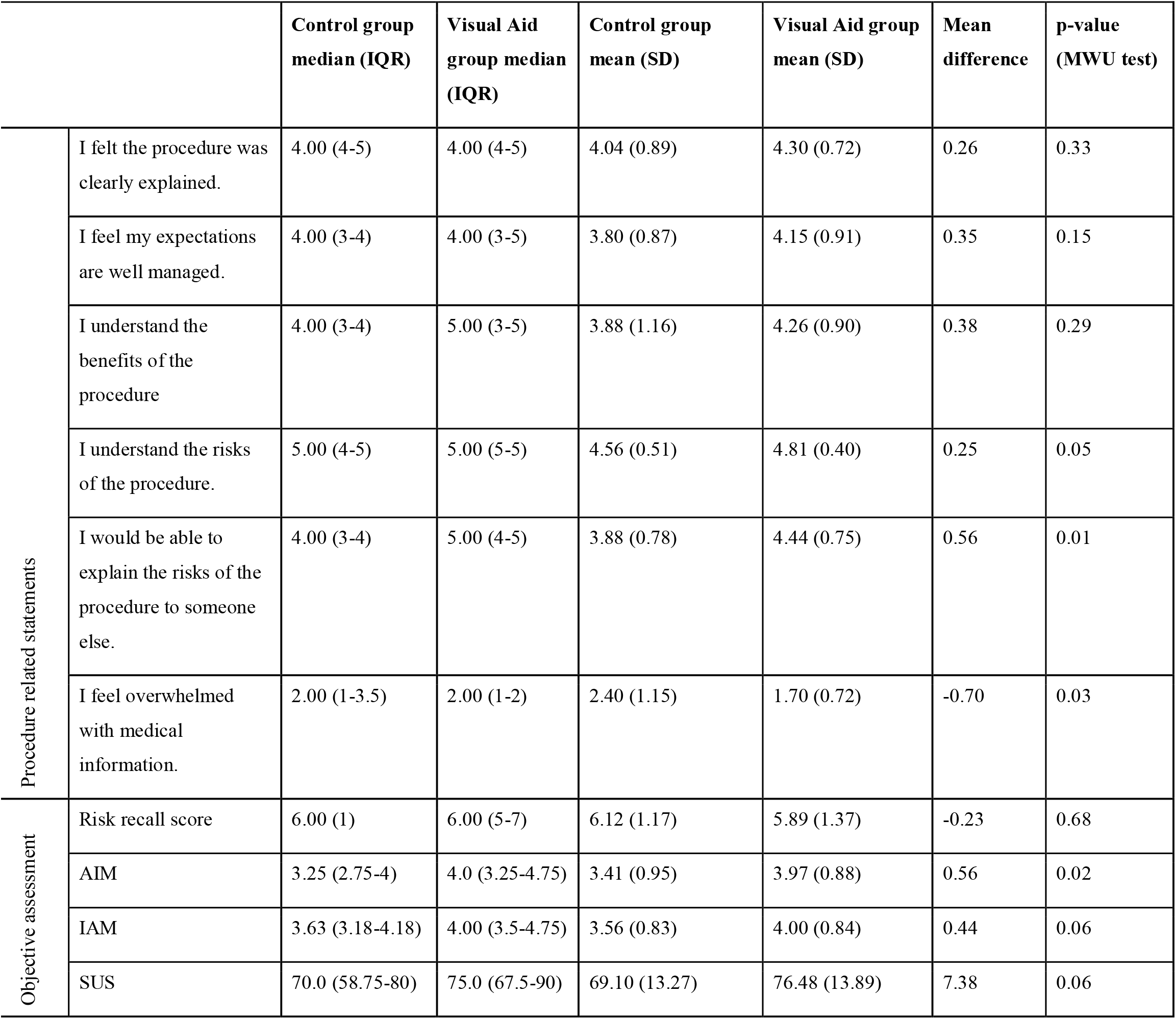
Outcomes by consent method. MWU = Mann-Whitney U test, IQR = Interquartile range, AIM = Acceptability of Intervention Measure, IAM = Intervention Appropriateness Measure, SUS = System Usability Scale

|  |  | Control group<br>median (IQR) | Visual Aid<br>group median<br>(IQR) | Control group<br>mean (SD) | Visual Aid group<br>mean (SD) | Mean<br>difference | p-value<br>(MWU test) |
| --- | --- | --- | --- | --- | --- | --- | --- |
| Procedure related statements | I felt the procedure was clearly explained. | 4.00 (4-5) | 4.00 (4-5) | 4.04 (0.89) | 4.30 (0.72) | 0.26 | 0.33 |
|  | I feel my expectations are well managed. | 4.00 (3-4) | 4.00 (3-5) | 3.80 (0.87) | 4.15 (0.91) | 0.35 | 0.15 |
|  | I understand the benefits of the procedure | 4.00 (3-4) | 5.00 (3-5) | 3.88 (1.16) | 4.26 (0.90) | 0.38 | 0.29 |
|  | I understand the risks of the procedure. | 5.00 (4-5) | 5.00 (5-5) | 4.56 (0.51) | 4.81 (0.40) | 0.25 | 0.05 |
|  | I would be able to explain the risks of the procedure to someone else. | 4.00 (3-4) | 5.00 (4-5) | 3.88 (0.78) | 4.44 (0.75) | 0.56 | 0.01 |
|  | I feel overwhelmed with medical information. | 2.00 (1-3.5) | 2.00 (1-2) | 2.40 (1.15) | 1.70 (0.72) | -0.70 | 0.03 |
| Objective assessment | Risk recall score | 6.00 (1) | 6.00 (5-7) | 6.12 (1.17) | 5.89 (1.37) | -0.23 | 0.68 |
|  | AIM | 3.25 (2.75-4) | 4.0 (3.25-4.75) | 3.41 (0.95) | 3.97 (0.88) | 0.56 | 0.02 |
|  | IAM | 3.63 (3.18-4.18) | 4.00 (3.5-4.75) | 3.56 (0.83) | 4.00 (0.84) | 0.44 | 0.06 |
|  | SUS | 70.0 (58.75-80) | 75.0 (67.5-90) | 69.10 (13.27) | 76.48 (13.89) | 7.38 | 0.06 |

## Discussion

Our pilot study suggests that that visual risk communication adjuncts may offer some advantages when compared to traditionally obtained surgical consent, particularly with reference to subjective understanding and attitudes toward, procedural risks. Statistically significant improvements were noted in the trial group regarding the ability to explain risks to others and greater acceptability and good usability of the consent adjunct, whilst also feeling less overloaded with medical information.

We acknowledge limitations in our approach, including the choice of individuals and the intervention. Recruiting healthy adults allowed us to test our hypothesis in a controlled simulated setting without bioethical concerns, but this does not fully replicate the atmosphere and anxieties associated with consent in hospital, nor having the procedure performed in the context of experiencing a disease process. The interventional choice of a lumbar puncture although simple remains a procedure that requires written informed consent and carries non-trivial complications (4). Further work should assess procedures of greater complexity and the impact of framing bias, namely, whether consent outcomes depend on the mode of presentation (5). Another limitation of the study is the overall high rate of literacy in both groups according to the table of demographics. In this case, the numerical literacy of the participants could be considered higher than expected in the general surgical population and therefore not easily generalisable.

This controlled trial evaluates the utility of risk communication adjuncts for surgical consent and contributes to this expanding field of bioethics. The surgical decision-making process is not only related to risk perception, but also to risk acceptance in accordance with an individual’s threshold for specific postoperative complications and the frequency in which they occur (6). To make consent fully informed, patient decision-making should be in line with, or adapted to, their probabilistic understanding of the intervention. Considering the findings of our study, we believe that visual risk communication tools could enable a more informed consent process. This pilot study provides a suggested methodology that could be useful in future larger-scale populations studies to replicate our conclusions.

## Data Availability

All data produced in the present study are available upon reasonable request to the authors

