## Supplementary Methods for "Improving risk communication: a pilot randomised-control trial assessing the impact of visual aids for interventional consent"

***Video Generation***

Please find the relevant video links below:

- **Group A (Control Group):** https://youtu.be/-QuOp-BTEKM" \h
- **Group B (Visual Aid Group):** https://youtu.be/wFe4KXEq8hw" \h

At the end of the videos, the participants of both groups were presented with a written consent form as it is used in clinical practice summarising the name, the risks and the benefits of the procedure. It was requested that participants sign the consent form 1 as they would do in clinical practice.

***Questionnaire Generation***

Participant knowledge and feedback with the respective consent process were assessed using a 41-item questionnaire developed in Qualtrics (Qualtrics, Provo, UT), the different sections and their respective questions are detailed in **Table 2**. The first section consisted of demographic information including age, gender and familiarity with the procedure. The second section required participants to rate their agreement to a series of seven statements specifically written for the study using a 5-point Likert scale. The intervention group received an additional three statements relating to the helpfulness of the three visual aids used in the video (Anatomy diagram, ten-man diagram and relative risk diagram). The ten-man diagram and the relative risk diagram are based on the Paling diagram and Paling Palette respectively with the permission of the Risk Communication Institute (Figure 2 & Figure 3) (1). The A free-text prompt asking for additional feedback to improve the consent process was also included. This was followed by seven multiple choice questions assessing participants’ knowledge on the procedure. The final section consisted of 19 questions to assess the implementation of the process in clinical practice using four validated scales (**Table 2**) (2-3).

| **Validated Scale** | **Description** | **Questions included** |
| --- | --- | --- |
| Procedure related questions | This section was composed of seven Likert scale statements (“Strongly disagree” to “Strongly agree) and was used to assess participant’s feelings towards their respective consent processes. The following statements were designed specifically for this study. | 1. I felt the procedure was clearly explained 2. I understand the benefits of the procedure 3. I understand the risks of the procedure 4. I feel my expectations are well managed 5. I would be able to explain the risks of the procedure to someone else 6. I feel overwhelmed with medical information |
| Knowledge Assessment | Questions were based on the information provided to both groups in each video and used to assess participant knowledge/recall of the procedure and its associated risks. The answers to each question were present in both videos. | 1. The needle for lumbar puncture is inserted in the    a. Spinal cord    b. Spinal canal    c. Subcutaneous tissue 2. The most common risk of lumbar puncture is    a. Headache    b. Back pain    c. Bleeding    d. Infection 3. What is the risk of infection to skin, brain or spinal cord?    a. More than 1 in 10,000    b. Equal to 1 in 10,000    c. Less than 1 in 10,000 4. What is the risk of a headache following a lumbar puncture?    a. 10%    b. 30%    c. 40% 5. What is the risk of back pain following a lumbar puncture?    a. 10%    b. 15%    c. 20% 6. What is the lifetime risk of death in the UK from a road traffic accident?    a. Approximately 42 in 10,000    b. Approximately 48 in 10,000    c. Approximately 52 in 10,000 7. What is the risk of bleeding following a lumbar puncture?    a. Less than 1%    b. Less than 2%    c. Less than 5% |
| The System Usability Score (SUS)^1^ | A reliable tool for measuring the usability of a system, a product or a method. It consists of a 10 item questionnaire, where the individuals provide responses ranging from “Strongly Agree” to “Strongly Disagree”. | 1) I think that, if I needed to, I would like to use this consent process 2) I found the consent process unnecessarily complex 3) I think that the way the risk was presented for this consent process was appealing 4) I think that I would need the support of a technical person to be able to use the consent process 5) I found the various functions in the consent process were well integrated 6) I thought there was too much inconsistency in the consent process 7) I would imagine that most people would learn to use this consent process very quickly 8) I found the consent process very cumbersome of use 9) I felt very confident using this consent process 10) I needed to learn a lot of things before I could get going with this consent process |
| Acceptability of Intervention Measure (AIM)^2^ | Acceptability is the perception among individuals that a given method is agreeable and satisfactory.  A Scale was created for each group and the average score of the following responses was recorded. The value of each response ranged from 1 to 5. | 1) This consent process meets my approval 2) This consent process is appealing to me 3) I like this consent process 4) I welcome this consent process |
| Intervention Appropriateness Measure (IAM)^2^ | Appropriateness is the perceived relevance and compatibility of the proposed method for consent among participants.  A Scale was created for each group and the average score of the following responses was recorded. The value of each response ranged from 1 to 5. | 1) This consent process is fitting 2) This consent process is suitable 3) This consent process is applicable 4) This consent process seems like a good match |

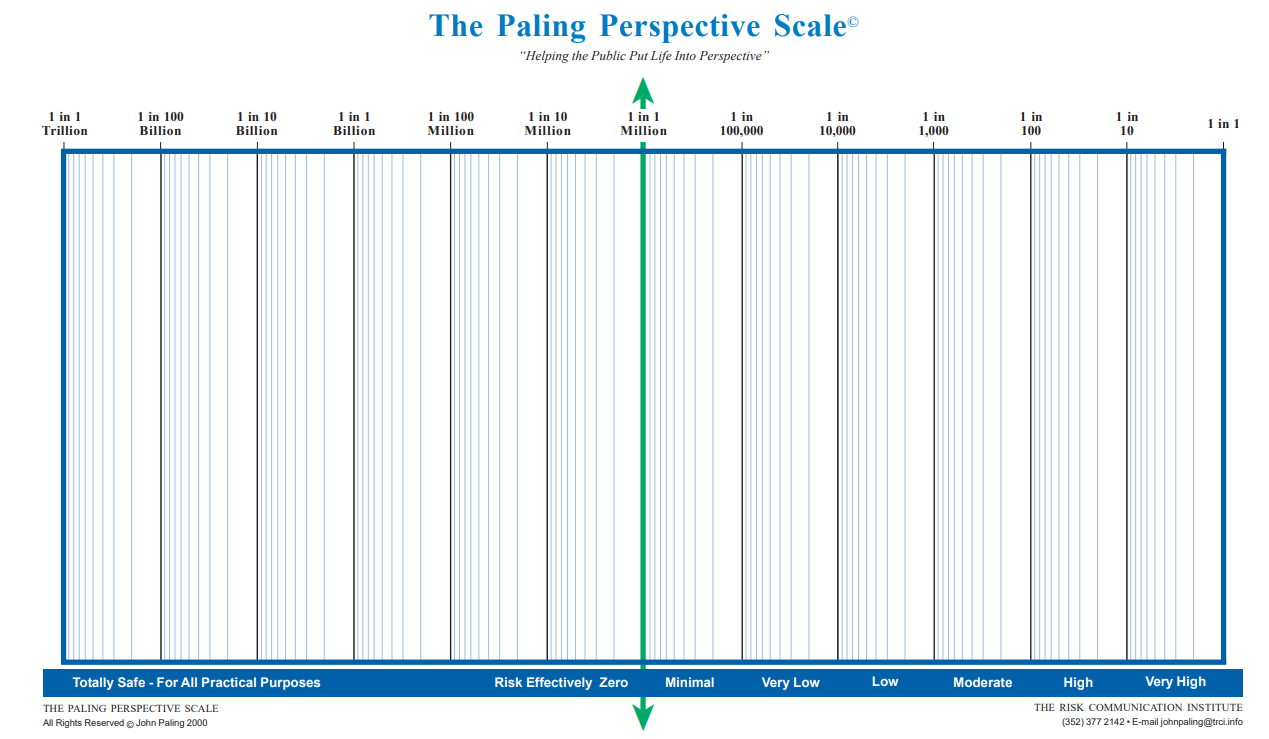

*Supplementary Figure 1. The Paling Perspective Scale. Reproduced with permission from The Risk Communication Institute*

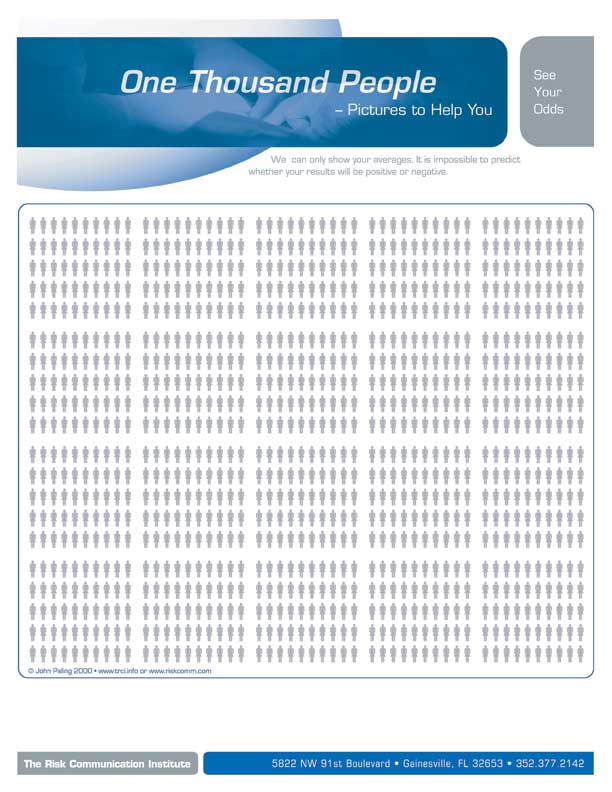

*Supplementary Figure 2. One Thousand People Figure. Reproduced with permission from The Risk Communication Institute*

***Participant recruitment***

Inclusion criteria were healthy individuals above 18 years old without any underlying cognitive impairment. Exclusion criteria were individuals with prior experience receiving, performing or observing the procedure, individuals lacking capacity to consent and hospitalised individuals. The study was advertised online via social media and mailing lists within our institutional academic community. Upon clicking the link to the questionnaire, participants were randomised in a 1:1 ratio using Qualtrics’ built in randomization software to receive either the control group with the standard informed consent proves or the intervention group which featured various visual aids. Researchers were blinded to the assignment of individual participants to the respective groups. Participant recruitment took place over a two-month period from 27^th^ March 2022 to 26^th^ May 2022. Data collection was concluded when the target sample size was reached.

***Ethical approval***

This study’s protocol was reviewed and approved by an institutional ethics committee (UCL Research Ethics Committee Project ID: 21837/001). Prior to being able to complete the questionnaire, participants were asked to confirm they meet the inclusion criteria of the study and voluntarily consent to completing the study through Qualtrics. Participants were directed to a participant information sheet within the study. Names and contact information of participants was not collected.

***Supplementary References***

1. Paling J. Strategies to help patients understand risks. BMJ. 2003 Sep 27;327(7417):745-8.

2. Brooke J. SUS: A quick and dirty usability scale. *Usability Eval Ind*. 11/30 1995;189.

3. Weiner BJ, Lewis CC, Stanick C, et al. Psychometric assessment of three newly developed implementation outcome measures. *Implement Sci*. Aug 29 2017;12(1):108.
